# A transcriptome-wide Mendelian randomization study in isolated human immune cells highlights risk genes involved in viral infections and potential drug repurposing opportunities for schizophrenia

**DOI:** 10.1101/2024.02.18.24303002

**Authors:** David Stacey, Liam Gaziano, Preethi Eldi, Catherine Toben, Beben Benyamin, S Hong Lee, Elina Hyppönen

## Abstract

**Background:** Schizophrenia is a neurodevelopmental psychiatric disorder characterised by symptoms of psychosis, thought disorder, and flattened affect. Immune mechanisms are associated with schizophrenia, though the precise nature of this relationship (i.e., causal, correlated, consequential) and the mechanisms involved are not fully understood.

**Methods:** To elucidate these mechanisms, we conducted a transcriptome-wide Mendelian randomization study using gene expression exposures from 29 human *cis*-eQTL datasets encompassing 11 unique immune cell types, all publicly available from the eQTL catalogue.

**Results:** These analyses highlighted 196 genes, including 67 located within the human leukocyte antigen (HLA) region. Enrichment analyses indicated an over-representation of immune genes, which was driven by the HLA genes. Stringent validation and replication steps retained 61 candidate genes, 27 of which were the sole causal signals at their respective loci, thereby representing strong candidate effector genes at known risk loci. We highlighted *L3HYPDH* as a potential novel schizophrenia risk gene and *DPYD* and *MAPK3* as candidate drug repurposing targets. Futhermore, we performed follow-up analyses focused on one of the candidate effectors, interferon regulatory transcription factor 3 (*IRF3*), which coordinates interferon responses to viral infections. We found evidence of shared genetic aetiology between schizophrenia and autoimmune diseases at the *IRF3* locus, and a significant enrichment of IRF3 chromatin binding at known schizophrenia risk loci.

**Conclusions:** Our findings highlight a novel schizophrenia risk gene, potential drug repurposing opportunities, and provide support for *IRF3* as a schizophrenia hub gene, which may play critical roles in mediating schizophrenia-autoimmune comorbidities and the impact of viral infections on schizophrenia risk.

## INTRODUCTION

Schizophrenia (SCZ) is a devastating psychiatric illness affecting approximately 1 in 300 people worldwide(1), with onset generally occurring during late adolescence or early adulthood(2). It is characterised by several debilitating symptoms including psychosis (i.e., hallucinations, delusions), disorganised thought and speech, and flattened affect(3). Although SCZ is highly heritable (∼80%)(4), its underlying genetic aetiology is complex and heterogeneous. Moreover, approximately 30% of patients are resistant to currently available antipsychotics(5), which underscores the need to identify novel drug targets. Importantly, recent studies have shown that anti-inflammatory medications can alleviate SCZ symptoms in some patients, including treatment-resistant patients(6, 7).

Immune hypotheses of SCZ have been in existence for decades, supported by several lines of evidence. For example, some people with SCZ have abnormalities in the levels of circulating immune markers(8, 9) and autoantibodies(10). People with SCZ also have an increased risk of comorbid immune-mediated diseases, particularly autoimmune diseases(11); while inflammation of the brain – or encephalitis – can often present with psychiatric symptoms such as psychosis(12). Although the relationship between the immune system and SCZ is highly complex and likely confounded by several lifestyle factors (e.g., smoking) and medication effects (e.g., antipsychotics), findings from Mendelian randomization (MR) studies, which are robust to such confounds, suggest that several circulating immune markers may be causally associated with SCZ risk(13–15).

Of these immune markers, C-reactive protein (CRP) – a key modulator of the complement cascade – has received the most widespread attention, with several independent MR studies consistently showing that a genetic propensity to higher levels of circulating CRP is associated with a lower risk of SCZ(13, 15, 16). The precise mechanisms underlying this association are currently unclear, though they may be mediated by protection against infection(17). Indeed, early life or maternal viral/bacterial infection is a well-established risk factor not only for SCZ(18, 19) but also autoimmune diseases(20), which may explain a proportion of the observed SCZ-autoimmune comorbidities.

Over the past decade, GWASes have identified more than 200 independent risk loci for SCZ, of which the most highly significant (by many orders of magnitude) resides within the human leukocyte antigen (HLA) region(21). Fine-mapping studies have highlighted *C4A*, which encodes complement component 4a, as a candidate effector gene at this locus, potentially mediated by perturbed synaptic pruning during adolescence(22). Nevertheless, pinpointing the precise effector genes at complex disease risk loci is a non-trivial task, and so the extent to which other immune-related genes may mediate some of the known genetic risk is currently unclear. The continued integration of GWAS data with functional genomic data from immune tissues or cells may help to address this knowledge gap.

To this end, we performed a transcriptome-wide MR (TWMR) study of SCZ using publicly available expression quantitative trait locus (eQTL) data from 11 isolated immune cell types of both myeloid and lymphoid origin, including microglia. The TWMR approach utilises *cis*-eQTLs to instrument gene expression exposures at scale, and is a proven method for identifying causal genes for complex human diseases(23, 24). We identified several immune-related genes as causal candidates for SCZ, potentially by modulating immune responses to neurotropic viral infection. We performed follow-up analyses of one of the immune gene candidates, interferon regulatory factor 3 (*IRF3*), which encodes a transcription factor responsible for inducing interferon responses to viral infections. We show that SCZ shares genetic aetiology with autoimmune diseases at the *IRF3* locus, and that IRF3 chromatin binding sites are significantly enriched at SCZ risk loci. We propose *IRF3* to be a SCZ hub gene and a potentially druggable therapeutic target.

## METHODS

### *Cis*-eQTL and GWAS summary datasets

We extracted 29 *cis*-eQTL datasets derived from isolated human immune cells from the eQTL catalogue (**Table 1**) (25, 26). Overall, these datasets represented 11 unique immune cell types. We extracted summary data from the largest and most recently published (at the time of writing) SCZ GWAS(21) from the psychiatric genetics consortium (PGC) (https://pgc.unc.edu/for-researchers/download-results/), which included up to 130,644 Europeans (53,386 cases). See **Supplementary Information** for a complete list of datasets we used and their sources.

**Table 1.** Summary of human immune cell *cis*-eQTL datasets downloaded from the eQTL catalogue for transcriptome-wide Mendelian randomization and statistical colocalization analyses.

| Study | Cell types | Conditions | Samples | Gene expression platform |
| --- | --- | --- | --- | --- |
| Alasoo et al. (2018)(38) | Macrophages | Naïve, IFN-γ, salmonella, IFN-γ & salmonella | 84 | RNA-seq |
| Blueprint(39) | Monocytes, Neutrophils, CD4+ T cells | Naïve | 197 | RNA-seq |
| Bossini-Castillo et al. (2019)(40) | Regulatory T cells | Naïve | 119 | RNA-seq |
| CAP(41) | LCLs | Naïve, statins | 148 | RNA-seq |
| CEDAR(42) | CD4+ T cells, CD8+ T cells, Monocytes, Neutrophils, B cells | Naïve | 322 | Microarray |
| Fairfax et al. (2012)(43) | B cells | Naïve | 282 | Microarray |
| Fairfax et al. (2014)(44) | Monocytes | Naïve, IFN-γ (24hrs), LPS (2hrs), LPS (24hrs) | 424 | Microarray |
| Gencord(45) | LCLs, T cells | Naïve | 195 | RNA-seq |
| GEUVADIS(46) | LCLs | Naïve | 445 | RNA-seq |
| Gilchrist et al. (2021)(47) | Natural Killer cells | Naïve | 247 | Microarray |
| Kasela et al. (2017)(48) | CD4+ T cells, CD8+ T cells | Naïve | 297 | Microarray |
| Naranbhai et al. (2015)(49) | Neutrophils | Naïve | 93 | Microarray |
| TwinsUK(50) | LCLs | Naïve | 433 | RNA-seq |
| Young et al. (2019)(51) | Microglia | Naïve | 104 | RNA-seq |

### Transcriptome-wide Mendelian randomization

We performed TWMR analyses using GSMR(24), as implemented in the Genome-Wide Complex Trait Analysis (GCTA) suite of tools (v1.94.1)(27). Prior to running GSMR, we first extracted all *cis*-eQTLs from the eQTL catalogue(25, 26) datasets with *p*-values lower than the estimated empirical *p*-values (calculated from 1,000 permutations) for the corresponding eGene based on the beta distribution. We obtained all pre-computed empirical *p*-values from the eQTL catalogue(25, 26). We supplied the pre-filtered exposure input files to GSMR and artificially set the GSMR significance threshold to *p*<0.05 (--gwas-thresh 0.05) to ensure all extracted *cis*-eQTLs were available for LD clumping within the GSMR workflow. Because *cis*-eQTL data generally yields fewer independent genetic instruments than well-powered GWASes, we set the minimum number of significant variants required for GSMR analysis to 1 (--gsmr-snp-min 1) and we applied a lenient LD (r^2^) threshold of 0.1 (--clump-r2 0.1) (GSMR natively adjusts estimates for correlated instruments). We set all other GSMR parameters to default.

To estimate variant correlations, we utilized an individual-level reference genotype panel comprising 4,994 participants from the INTERVAL population study(28). To minimise type I errors, we adjusted significance thresholds by applying a Bonferroni correction per eQTL dataset for the total number of eGenes tested (i.e., 0.05 / *n* eGenes). We plotted the GSMR estimates using the forestplot (v3.1.1) (https://CRAN.R-project.org/package=forestplot) and corrplot (v0.92)(29) R packages.

### Statistical colocalization

We performed statistical colocalization analyses using two software tools: (i) the Coloc R package (v5.1.0.1)(30); and (ii) a C++ implementation of pair-wise conditional and colocalization (PWCoCo) analysis(31). Coloc and PWCoCo have been described previously, but for brief descriptions, see **Supplementary Information**.

### Gene ontology term enrichment analyses

We conducted gene ontology (GO) term enrichment analyses using the functional annotation chart webtool available at the Database for Annotation, Visualization, and Integrated Discovery (DAVID)(32). To facilitate interpretation, we performed enrichment analyses focused solely on the gene ontology (GO) direct terms for biological process, molecular function, and cellular component. We unselected all other functional annotation sources before beginning analyses. For the background gene set, we extracted all unique eGenes across all eQTL catalogue datasets (**Table 1**) used in TWMR analyses.

### Phenome-wide association studies

We conducted single variant phenome-wide association studies (PheWASs) using the Open Targets Genetics(33) and the integrated epidemiology unit (IEU) OpenGWAS(34) platforms. To search the Open Targets Genetics platform, we used the webtool. For each variant of interest, we extracted all phenotypes from the ‘GWAS lead variants’ module, which contained phenotypes/studies with a lead variant linked with the variant of interest either through linkage disequilibrium (r^2^>0.5) or fine-mapping (i.e., appears in the 95% credible causal set). To search the IEU OpenGWAS platform, we used the ieugwasr R package (v0.1.5) with the *p*-value threshold set to *p*<5×10^-8^.

### Distance-based clumping

We ordered the 61 SCZ-associated variants by chromosome and position, and extracted all genes from the xHLA region (chr6:25,726,063-33,400,644; hg38)(35). We first assigned all xHLA region genes to a single mega-clump. For the remaining non-xHLA genes, we used the first gene to initiate the first clump and then sequentially checked the distance between the next gene and the preceding one. If the distance was <500Kb, we added the gene to the current clump; and if the distance was ≥500Kb, we used that gene to initiate the next clump. We continued this process until all genes were assigned to a clump.

### Enrichment analysis of regulatory data

To determine whether IRF3 ChIP-seq peaks were enriched at SCZ risk loci, we utilized the GWAS analysis of regulatory or functional information enrichment with LD correction (GARFIELD) v2 software tool(36), available at https://www.ebi.ac.uk/birney-srv/GARFIELD/. For further information, see **Supplementary Information**.

## RESULTS

### Transcriptome-wide Mendelian randomization (TWMR) analyses of isolated human immune cell types highlights 196 candidate schizophrenia-associated genes

**Fig. 1** describes the overall study design and main analyses conducted. To identify immune genes with potential causal roles in SCZ, we performed TWMR analyses using summary data from 29 *cis*-eQTL datasets selected (see **Methods**) from the eQTL catalogue generated by 15 different studies from 11 unique immune cell types (**Table 1**). To facilitate future TWMR efforts focused on exposures from immune cells, we have provided as an immunogenomic resource all the genetic variants used to instrument genes in each of the eQTL catalogue datasets we tested (**Table S1**). Overall, these analyses highlighted a total of 196 significant genes (see **Methods, Fig. 2a, Table S2**), of which 156 (79.59%) were protein-coding and 67 (34.18%) were located within the extended HLA (xHLA) region as defined by Horton et al. (2004) (chr6:25,726,063-33,400,644; hg38)(35). Notably, this gene set did not include *CRP* or any genes encoding major cytokines. The absence of *CRP* was expected since it is specifically expressed in the liver, with only residual expression observed in immune cells(37).

**Figure 1.**
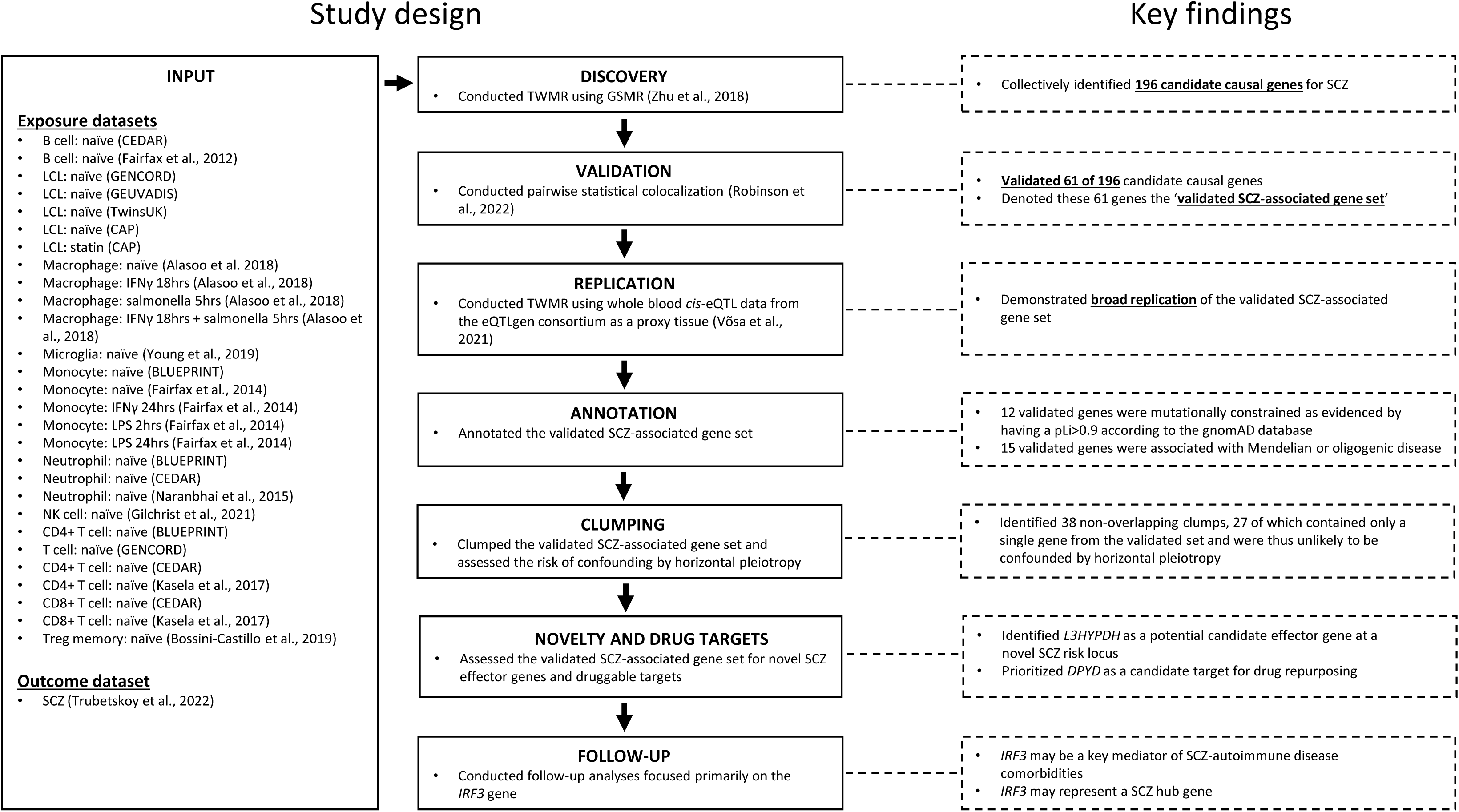
Flow diagram summarising the overall study design and key findings from each analysis step. GSMR: generalised summary-data-based Mendelian randomization, GTEx: genotype-tissue expression project, SCZ: schizophrenia, TWMR: transcriptome-wide Mendelian randomization; pLi: probability of loss of function intolerance.

**Figure 2.**
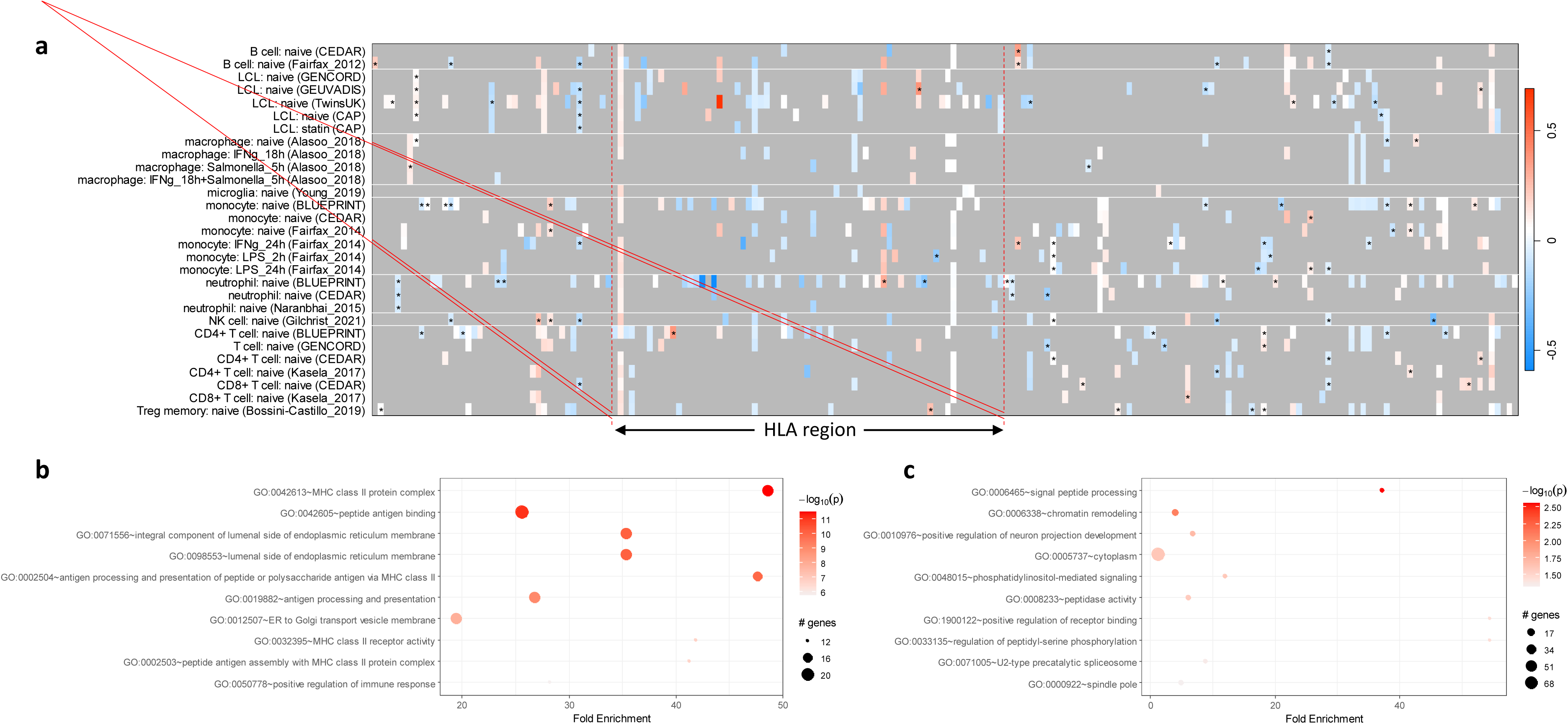
Transcriptome-wide Mendelian randomization analyses identifies schizophrenia-associated candidate genes. (a) Heatmap plotting the bxy (logOR) estimates of the causal effect of isolated immune cell gene expression exposures on schizophrenia risk (y-axis: *cis*-eQTL datasets; x-axis: genes). Gene expression exposures with robust evidence of colocalization (PP_H4_>0.8) with schizophrenia risk are shown with asterisks. Vertical red dashed lines indicate the location of the human leukocyte antigen region (HLA). (b,c) Gene ontology term enrichment analyses with (b) and without (c) the HLA region genes included.

To characterise this list of 196 genes, we performed gene ontology (GO) term enrichment analyses using the Database for Annotation, Visualization, and Integrated Discovery (DAVID) (see **Methods**)(32). An initial analysis including all 196 genes revealed significant (FDR<0.05) enrichment of 30 GO terms, most of which were immune-related (**Fig. 2b**, **Table S3**). However, none of these terms retained significance after removing the 67 HLA region genes from the test set (**Fig. 2c**, **Table S4**), demonstrating that the immune enrichment observed in our initial analysis was driven by the HLA region genes.

### Statistical colocalization analyses provide validation for 61 schizophrenia-associated genes, 27 of which are the sole causal signals at their respective loci

Since TWMR studies are susceptible to confounding by linkage disequilibrium (LD), we sought to validate our findings by performing statistical colocalization analyses (see **Methods**). Overall, we found robust evidence (PP_H4_>0.8) of colocalization between SCZ and at least one *cis*-eQTL dataset for 61 of 196 genes (31.12%) (**Fig. 2a**, **Fig. 3**, **Table S5**) – herein, we will refer to these colocalising genes as the validated SCZ-associated gene set.

**Figure 3.**
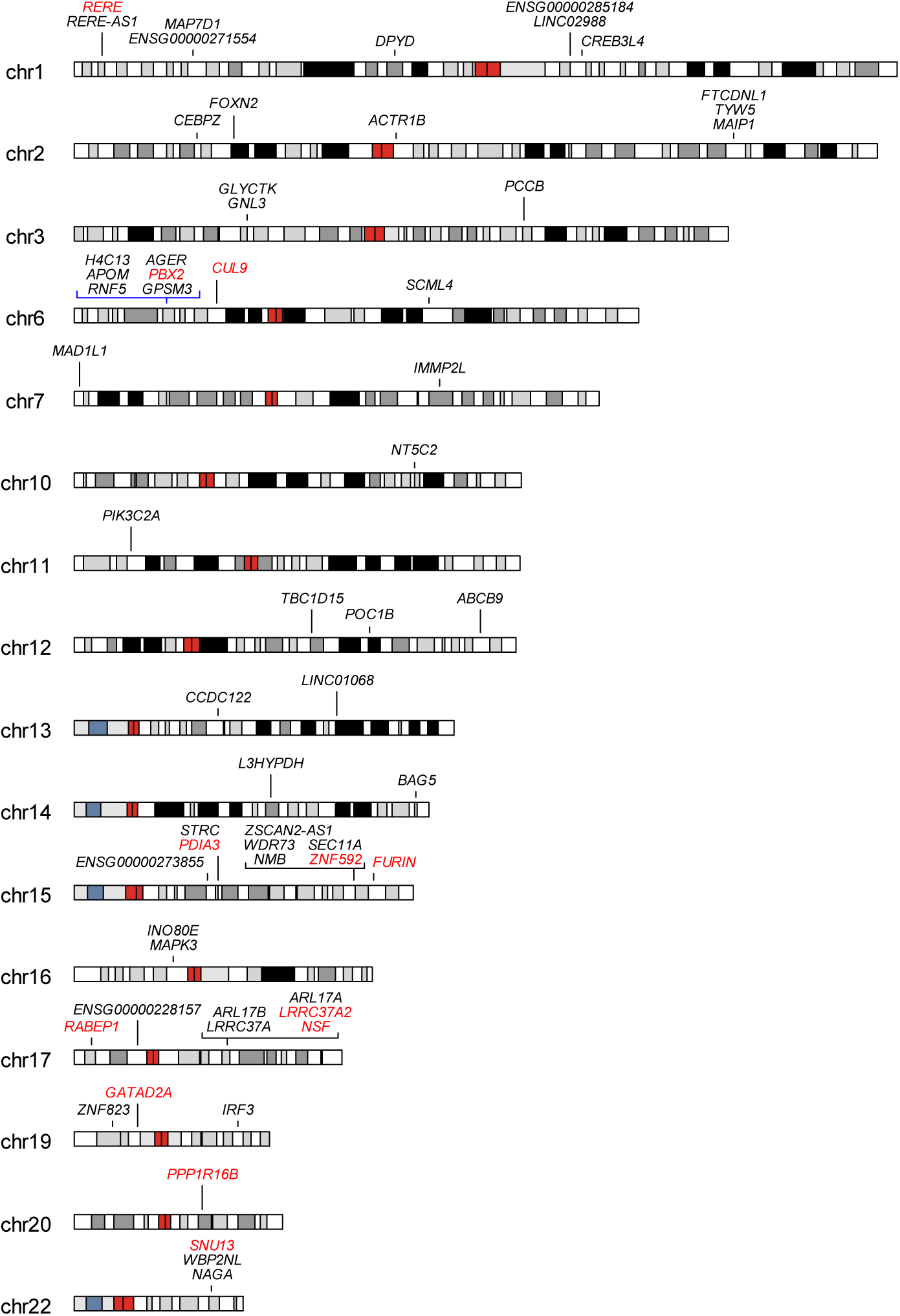
The validated SCZ-associated gene set contains candidate SCZ effector genes at 38 non-overlapping genomic regions. Ideogram of the human genome showing the locations of the 61 validated SCZ-associated gene sets. For brevity, chromosomes without a validated signal are not shown. The blue arrows indicate the extended human leukocyte antigen (xHLA) region, and genes with a probability of loss of function intolerance (pLi) > 0.9 are shown in red font.

Most of these 61 genes replicated in the whole blood *cis*-eQTL dataset from the eQTLgen consortium (**Fig. S1**, **Table** S6, see **Supplementary Information**), and many were mutationally constrained and associated with Mendelian or oligogenic diseases featuring neurodevelopmental, neurological, or immunological features (**Table S7**, see **Supplementary Information**). Moreover, most of the colocalizing genes resided outside of the extended HLA region, and the overall evidence for colocalization within the HLA region was sparse (**Fig. 2a**). Indeed, of the 67 xHLA region genes, only six of them (**Fig. 3**; *H4C13*, *APOM*, *RNF5*, *AGER*, *PBX2*, *GPSM3*) were in the validated SCZ-associated gene set, indicating that most of the associations observed in this region do not reflect causal effects on SCZ (at least not in the cell types tested here).

Horizontal pleiotropy is a major source of potential confounding in Mendelian randomization studies, occuring when an instrumental variable has an effect on the outcome outside of its effect on the exposure in question(52). Due to the extensive co-regulation of neighbouring genes, gene expression exposures can be particularly prone to horizontal pleiotropy(53). To assess this risk across our validated set of 61 SCZ-associated genes, we performed distance-based clumping using a 500kb window (see **Methods**). Including the xHLA region (chr6:25,726,063-33400644) as a single clump, this procedure revealed 38 non-overlapping clumps in total, 27 of which contained just a single gene from the validated SCZ-associated gene set (**Fig. 3**, **Table S8**). These 27 genes therefore represent high confidence effector genes for SCZ that are unlikely to be confounded by horizontal pleiotropy.

Of the remaining 11 bi- or multi-gene clumps, we were able to distinguish between genuine signals and those confounded by horizontal pleiotropy for three of them by manual curation (see **Supplementary Information**). This included the *NAGA*-*WBP2NL*-*SNU13* locus on chromosome 22, which contained two independent SCZ GWAS signals, one of which colocalized solely with *SNU13* mRNA expression in CD8+ T cells (**Fig. S2**).

### *L3HYPDH* is a candidate effector gene at a novel SCZ risk locus

Next, we sought to identify genes from the validated SCZ-associated set that did not reside at a genome-wide significant locus for SCZ, and may therefore represent novel SCZ risk genes. We intersected the genomic coordinates of the validated SCZ-associated gene set with the coordinates of the sentinel variants at the independent SCZ genome-wide significant loci reported by Trubetskoy et al. (2022), and found three genes that did not reside within 1Mb of a SCZ risk locus: *FOXN2*, *L3HYPDH*, *PIK3C2A*. We conducted look-ups in the Open Targets Genetics(33) and the IEU OpenGWAS(34) platforms to check whether any of these three genes or loci had been implicated in SCZ by any previous GWASs (see **Supplementary Results**, **Table S9**, **Table S10**). These look-ups revealed that the *FOXN2* and *PIK3C2A* signals, but not the *L3HYPDH* signal, had been detected in previous SCZ GWASs (see **Supplementary Results**, **Fig. S3**, **Table S11**, **Table S12**). Taken together, these findings suggest that *L3HYPDH* is a candidate effector gene at a novel SCZ risk locus.

### The schizophrenia-associated genes *DPYD* and *MAPK3* may present opportunities for drug repurposing

To identify potentially druggable targets of any of the proteins encoded by the 61 validated SCZ-associated genes, we performed gene-based look ups using the online DrugBank resource(54). This search revealed that the products of five of the genes (*DPYD*, *MAPK3*, *PCCB*, *NT5C2*, *FURIN*) were potentially druggable (**Table S13**).

The most robust evidence was for the dihydropyrimidine dehydrogenase (DPD) (*DPYD*) enzyme, which was directly and specifically targeted by a clinically approved drug called tegafur-uracil (**Table S13**). Tegafur-uracil is a chemotherapy drug combination primarily used in the treatment of bowel cancer. It comprises the prodrug tegafur, which is metabolised into 5-fluorouracil (5-FU) and incorporated into the DNA of dividing cancer cells, thereby killing them. However, since 5-FU is inactivated by DPD, tegafur-uracil also comprises an excess of uracil, which prolongs the half-life of 5-FU by competitively binding to DPD. As an alternative to uracil, tegafur is often combined with more potent inhibitors of DPD such as gimeracil. In our study, TWMR analyses indicated that higher expression levels of *DPYD* in a colocalizing (PP_H4_>0.8) eQTL macrophage (salmonella-treated) dataset(55) was associated with a higher risk of SCZ (β=0.120, se=0.026, *p*=3.60×10^-6^), and so compounds acting to reduce the activity of DPD like uracil and gimeracil may represent feasible therapeutic candidates.

Additionally, three approved compounds (arsenic trioxide, sulindac, cholecystokinin) are known to target the mitogen-activated protein kinase 3 (MAPK3), albeit with less specificity than the DPD-targeting compounds (**Table S13**). Most notably, sulindac, which inhibits MAPK3, is a nonsteroidal anti-inflammatory drug (NSAID) used in the treatment of acute and chronic inflammatory conditions. In our study, TWMR analyses indicated that higher *MAPK3* mRNA expression in two distinct colocalizing (PP_H4_>0.8) monocyte (naïve and LPS-treated) *cis*-eQTL datasets(42, 44) was associated with a higher risk of SCZ (naïve: β=0.229, se=0.046, *p*=5.29×10^-7^; LPS-treated: β=0.108, se=0.022, *p*=9.72×10^-7^), suggesting that sulindac may represent an opportunity for immune drug repurposing. Moreover, there may be other opportunities for the repurposing of immune drugs that have not yet been approved. For example, taribavirin and seliciclib, which target cytosolic purine 5’-nucleotidase (*NT5C2*) and MAPK3, respectively, are both currently under investigation for their efficacy in treating viral infections including hepatitis C and herpes simplex virus (**Table S13**).

### Interferon regulatory factor 3 is a schizophrenia hub gene and may contribute towards schizophrenia-autoimmune comorbidities

Next, we sought to further elucidate the immune component of SCZ by performing follow-up analyses focused on one of the genes from the validated SCZ-associated gene set. We selected the *IRF3* gene because it is a strong candidate effector gene at a robust genome-wide significant locus for SCZ on chromosome 19 (**Fig. 4a**)(21) and a well-established immune gene. *IRF3* encodes interferon regulatory factor 3, which is a transcription factor responsible for orchestrating interferon responses to viral infections(56). Moreover, rare loss of function variants affecting *IRF3* are associated with HSV-1-induced encephalitis(57), which is characterised by central inflammation and can present with neuropsychiatric symptoms including psychosis (**Table S7**)(12). In our TWMR analyses, we found an association between lower *IRF3* mRNA expression in natural killer (NK) cells(47) and a higher risk of SCZ (**Fig. 4a**,**b**; bxy= -0.32, se=0.07, *p*=5.96×10^-6^), and we also observed directionally consistent sub-threshold associations in both B cells(43) and neutrophils(39) (**Fig. 4b**).

**Figure 4.**
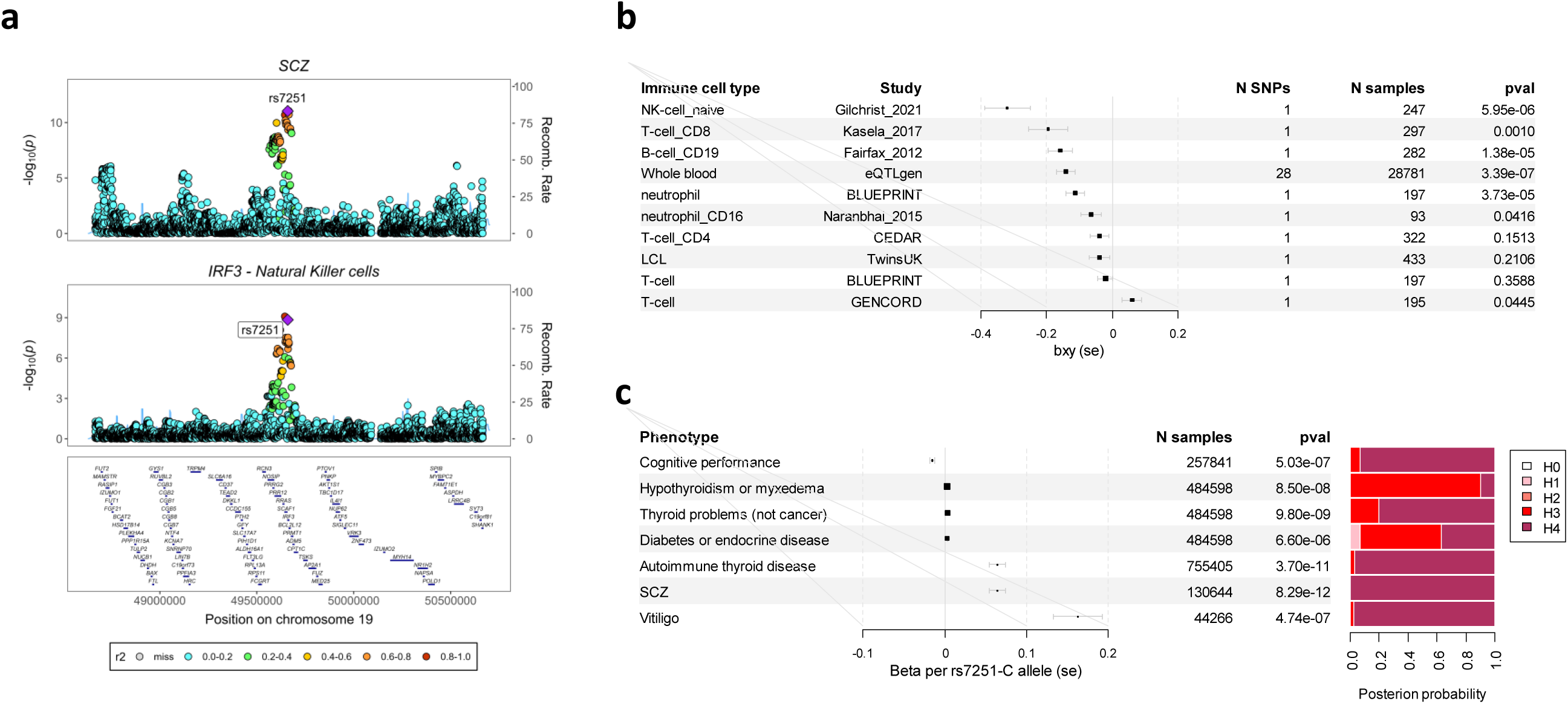
The rs7251 schizophrenia risk locus is associated with peripheral and central *IRF3* mRNA expression as well as risk of autoimmune diseases. (a) Forest plot depicting the bxy (logOR) estimates of the causal effect of immune cell and whole blood *IRF3* mRNA expression on schizophrenia risk. (b) Stacked regional association plot showing the genetic association signals at the *IRF3* locus for schizophrenia (top) and *IRF3* mRNA expression in natural killer cells (bottom). (c) Forest plot showing the results from a PheWAS of rs7251 and a stacked bar plot depicting PPs of colocalization between schizophrenia and each rs7251-associated phenotype.

To identify additional diseases and traits that might be associated with the *IRF3* locus, we performed a PheWAS of rs7251 using the Open Targets Genetics(33) platform (see **Methods**). This revealed six genome-wide significant associations (*p*<5×10^-8^) with either rs7251 or a proxy, most of which were autoimmune diseases involving thyroid dysfunction (**Fig. 4c**). Importantly, the same allele (rs7251-C) was associated with a higher risk of both SCZ and autoimmune disease, as well as poorer cognitive performance (**Fig. 4c**). We found robust evidence of colocalization (PP_H4_>0.8) with SCZ for four of the six phenotypes: autoimmune thyroid disease, cognitive performance, thyroid problems (not cancer), and vitiligo (an autoimmune disease affecting the skin) (**Fig. 4c**). These findings suggest that *IRF3* may contribute towards the observed comorbidity between SCZ and autoimmune diseases, particularly those involving thyroid dysfunction.

Transcription factors are often hub genes responsible for orchestrating complex transcriptional networks that regulate important biological processes. Since *IRF3* is a transcription factor, we sought to assess the extent to which *IRF3* might regulate the expression of other SCZ risk genes. To explore this, we performed enrichment analyses using GARFIELD(36) (GWAS analysis of regulatory or functional enrichment with LD correction) by leveraging (i) full SCZ GWAS summary data from Trubetskoy et al. (2022)(21) and (ii) all available chromatin immunoprecipitation sequence (ChIP-seq) data for IRF3 from the ENCODE project(58) (see **Methods**). We found significant enrichment of SCZ risk loci (defined using genome-wide [*p*<5×10^-8^] and suggestive [*p*<1×10^-5^] significance thresholds) at IRF3 peaks from lymphoblastoid, epithelial, and neuroblastoma cells (**Fig. 5a**).

**Figure 5.**
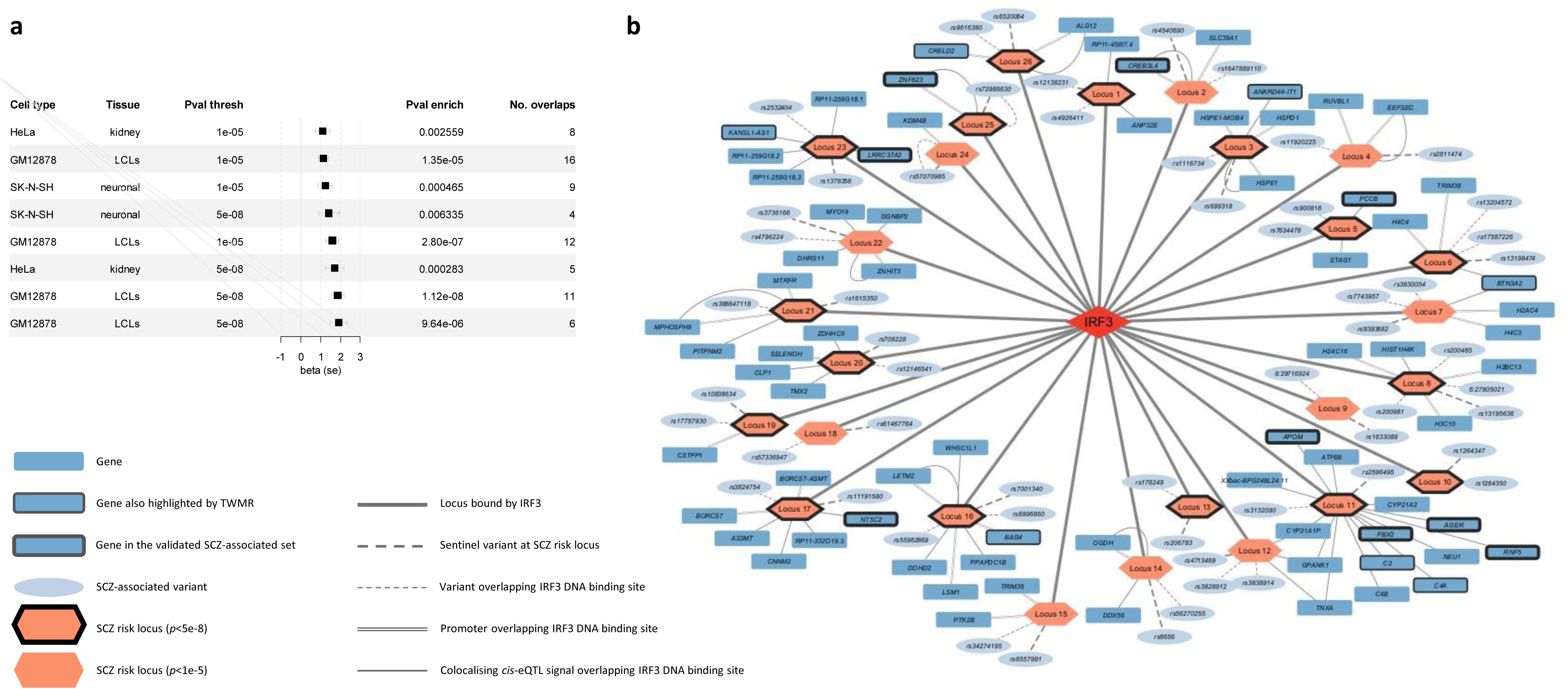
Interferon regulatory transcription factor 3 (IRF3) DNA binding sites are enriched at known schizophrenia risk loci. (a) Forest plot showing beta estimates (se) from GARFIELD enrichment analyses. (b) Graph showing the 26 schizophrenia risk loci with an overlapping IRF3 DNA-binding site as evidenced by chromatin immunoprecipitation-sequence (ChIP-seq) data. Colocalising (PP_H4_>0.8) *cis*-eQTL signals were from whole blood (eQTLgen consortium). SCZ: schizophrenia.

Overall, IRF3 peaks overlapped 15 independent genome-wide significant (*p*<5×10^-8^) and 11 suggestive (*p*<1×10^-5^) SCZ risk loci in at least one of the three significant cell types (**Fig. 5b**). We found that 18 of these loci (11 genome-wide significant, 7 suggestive) overlapped with IRF3 peaks at one or more gene promoters (+/-1Kb of the transcription start site), several of which were cognate to genes from our set of validated SCZ-associated genes (e.g., *ZNF823* and *CREB3L4*), suggesting *IRF3* may regulate the expression of these and other genes. Accordingly, we found robust evidence of colocalization between the SCZ signals at the 26 regions with whole blood *cis*-eQTL signals from eQTLgen for a total of 49 genes (**Table S14**, **Fig. 5b**). Many of these colocalizing *cis*-eQTL signals were cognate to the IRF3 peak-overlapping promoters, though not all of the, which suggests IRF3 may also be binding to other regulatory elements (e.g., enhancers) at some of these SCZ risk loci. Taken together, these findings are consistent with a model whereby SCZ-associated variants at these 26 risk loci exert regulatory effects on local gene expression either by strengthening or weakening IRF3 binding sites.

## DISCUSSION

In this TWMR study, we sought to identify immune genes and elucidate immune mechanisms involved in SCZ by leveraging 29 *cis*-eQTL datasets generated by 15 different studies from 11 unique human immune cell types, all publicly available from the eQTL catalogue(25, 26). Overall, TWMR analyses highlighted 196 unique candidate genes, 67 of which resided within the extended HLA region. We applied a series of stringent validation and replication steps culminating in the prioritization of 61 validated SCZ-associated genes (**Fig. 3**), 30 of which were unlikely to be confounded by horizontal pleiotropy and therefore represent convincing candidate SCZ effector genes. We highlighted *L3HYPDH* as a potentially novel SCZ risk gene and *DPYD* and *MAPK3* as candidate drug repurposing targets. We also conducted in-depth follow-up analyses of *IRF3*, showing it to be a candidate SCZ hub gene and a potential driver of SCZ-autoimmune comorbidities. Finally, we have made available as an immunogenomic resource all the genetic variants used in this study to instrument gene expression exposures from isolated human immune cells (**Table S1**). We hope this catalogue will facilitate future TWMR screening efforts interested in uncovering immune mechanisms of psychiatric disorders and other immune-mediated phenotypes.

A landmark study led by investigators at GlaxoSmithKline used retrospective data to show that drugs targeting genes and proteins supported by genetic evidence are more likely to progress through the drug development pipeline than those without genetic evidence(59). This has led to several large-scale MR studies investigating gene expression and protein abundance exposures seeking to identify novel drug targets and drug repurposing opportunities(60, 61). In this TWMR study, we found that the DPD (encoded by *DPYD*) inhibitor gimeracil, an adjunctive medicine used to enhance the efficacy of a cancer medicine called tegafur, may represent a drug repurposing opportunity for SCZ. DPD is an enzyme involved in the metabolism of the pyrimidines uracil and thymine, which function as building blocks of nucleic acids, play roles in energy metabolism, and can bind to P2Y receptors(62). Moreover, people with DPD deficiency present with – amongst other symptoms – intellectual disability and impaired social functioning(63), suggesting that gimeracil may help to treat SCZ symptoms relating to cognitive and social impairment.

As a means to further elucidate the immune component of SCZ, we conducted deep-dive analyses focused on *IRF3*, a well characterised immune gene from our validated SCZ-associated gene set. *IRF3* is a downstream effector of the cyclic GMP-AMP synthase (cGAS)-stimulator of interferon genes (STING) pathway, and is responsible for coordinating interferon type I responses following the detection of viral DNA by cGAS(64). Our findings indicated that a genetic propensity to lower *IRF3* expression was associated with a higher risk of SCZ (**Fig. 4b**), which would be consistent with a blunted interferon response to viral infections as a potential SCZ risk factor. A blunted interferon response could lead to the incomplete resolution of viral infections, thereby promoting viral latency and subsequent chronic intermittent or low-grade inflammation(65). Inflammatory mechanisms such as these, particularly those affecting the brain, have long been hypothesised to increase the risk of SCZ or SCZ-like symptoms.

Accordingly, rare loss of function variants affecting *IRF3* lead to HSV-1-induced encephalitis, which is characterised by central inflammation and can present with neuropsychiatric symptoms including psychosis(12, 57). HSV-1 is one of several viruses implicated in the pathophysiology of SCZ, and it is a compelling candidate because it is neurotropic – that is, capable of infecting the brain. Indeed, HSV-1 initially infects epithelial cells at orofacial surfaces(66), after which it can travel afferently along sensory neurons from the skin into the brain and has been shown to specifically infect areas of the limbic system, where it can remain latent(67). Our findings implicated *IRF3* in NK cells, which are amongst the first responders to viral infections(68). We therefore propose a model whereby *IRF3* activity in NK cells might modulate SCZ risk by acting as an early buffering system protecting against the spread of neurotropic viral infections from the periphery into the brain. However, since *IRF3* is also expressed in the brain(69), we cannot rule out a direct brain-centric role for *IRF3* in modulating SCZ risk.

Notably, *IRF3* was not the only gene from the validated SCZ-associated gene set involved in the regulation of viral infections. *ZNF823* and *MAPK3* are both annotated to the HSV-1 infection (hsa05168) KEGG pathway(70), while *MAPK3* (and *IRF3*) was also found to be up-regulated in placentas from pregnancies with SARS-CoV-2 (COVID-19) maternal infection(23). *RNF5* has been shown to inhibit type I interferon responses in HSV keratitis through the cGAS-STING/IRF3 signalling pathway(71), and *FURIN* encodes a protease responsible for cleaving proteins produced by several viruses, including herpesvirus glycoprotein B produced by HSV, as well as the SARS-CoV-2 (COVID-19) spike protein(72). Moreover, a recent study has shown that *L3HYPDH*, potentially a novel SCZ risk gene according to this TWMR study, is induced by type I interferon signalling and inhibits the replication of enterovirus 71 (EV71) by interfering with the translation of EV71 proteins(73). The recurrence of this viral regulation theme suggests that this TWMR study, which focused solely on gene expression exposures from isolated human immune cells, has uncovered genes and mechanisms responsible for mediating the relationship between viral infections and SCZ.

There are several potential limitations to this study. First, our TWMR analysis was conducted with exposures from nearly 30 *cis*-eQTL datasets, posing an increased risk of type I errors. To minimise this risk, we applied a conservative Bonferroni correction to each dataset, (see **Methods**) complemented by stringent validation and replication analyses. Second, most of the *cis*-eQTL datasets from isolated immune cells were derived from relatively small sample sizes, which limits discovery power. Access to larger datasets in the future will facilitate more robust discoveries. Third, certain immune cell types tested were represented by a single *cis*-eQTL study (e.g., NK cells, microglia). Independent *cis*-eQTL datasets for these cell types are necessary for replication and to further explore their role in SCZ.

In conclusion, by conducting MR analyses using gene expression exposures from 29 *cis*-eQTL immune cell datasets followed by stringent validation steps, we highlighted 61 candidate effector genes for SCZ. Overall, response to viral infections was a recurring theme of these genes, and so future studies focusing on them will be well-placed to further elucidate the critical mechanisms mediating the established relationship between viral infections and SCZ. We exemplified this by conducting follow-up analyses of *IRF3*, which encodes a transcription factor responsible for orchestrating interferon responses to viral infections(56), and showed it to be a candidate SCZ hub gene that also modulates risk of autoimmune disease. Based on this, we propose that addressing unmet needs in vaccination (e.g., HSV-1) and the use of antiviral drugs may represent a viable path towards mitigating SCZ risk in the population.

## ACKNOWLEDGEMENTS

We would like to thank Dr. Kaur Alasoo and colleagues for their excellent eQTL catalogue resource.

## DATA AVAILABILITY

All data generated by this study are available as Supplementary Tables or will be provided upon request from the corresponding author. All transcriptomic exposure data used in this study are publicly available either from the eQTL catalogue (https://www.ebi.ac.uk/eqtl/Data_access/) or the Genotype-Tissue Expression project (https://gtexportal.org/home/downloads/adult-gtex/overview). Full GWAS summary data for schizophrenia from Trubetskoy et al. (2022)(21) are publicly available from the Psychiatric Genomics Consortium (PGC) website (https://pgc.unc.edu/for-researchers/download-results/).

