## Supplementary figures and images for "A transcriptome-wide Mendelian randomization study in isolated human immune cells highlights risk genes involved in viral infections and potential drug repurposing opportunities for schizophrenia"

### Supplementary Figures

#### Slide 1
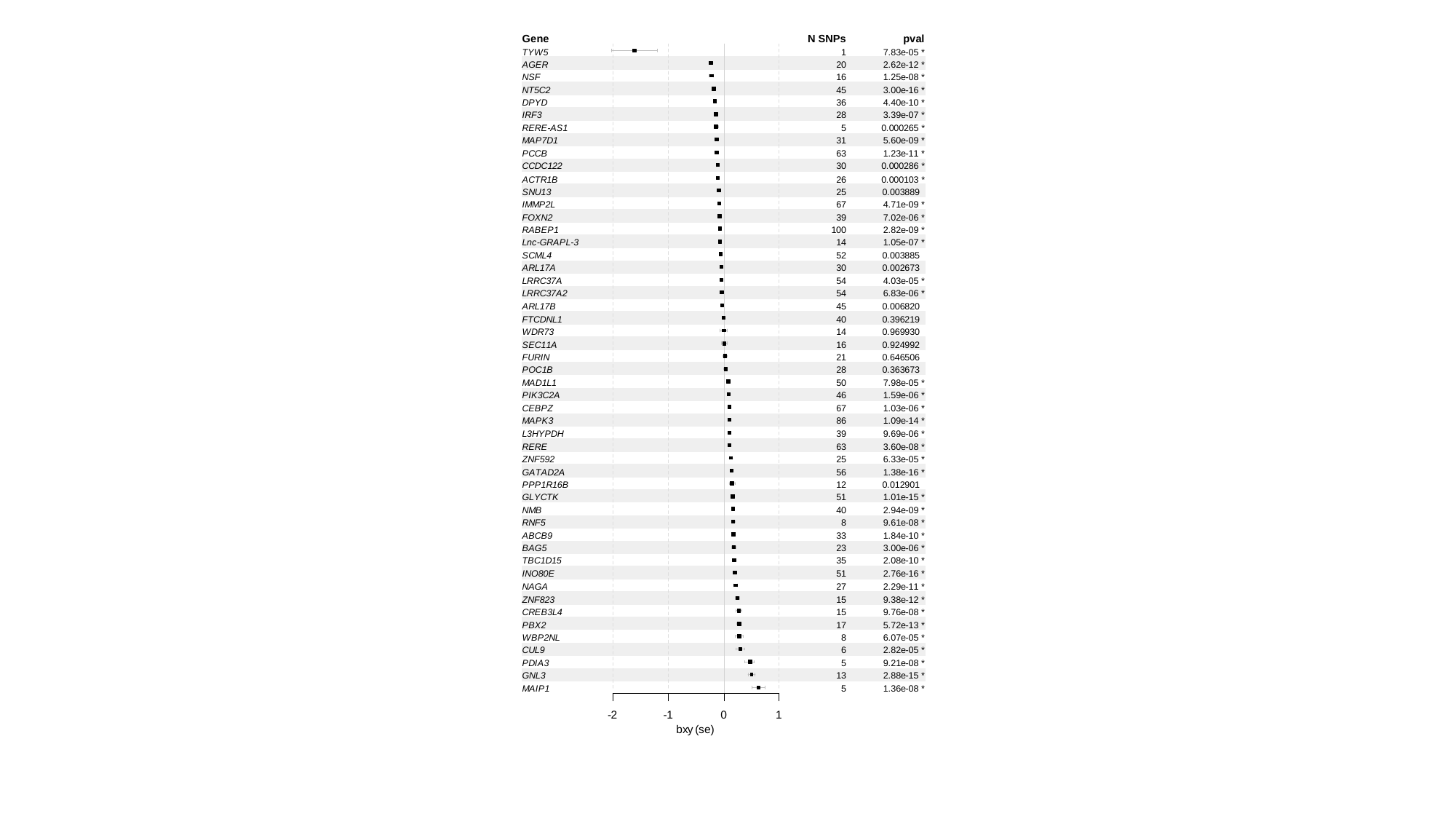

#### Slide 2
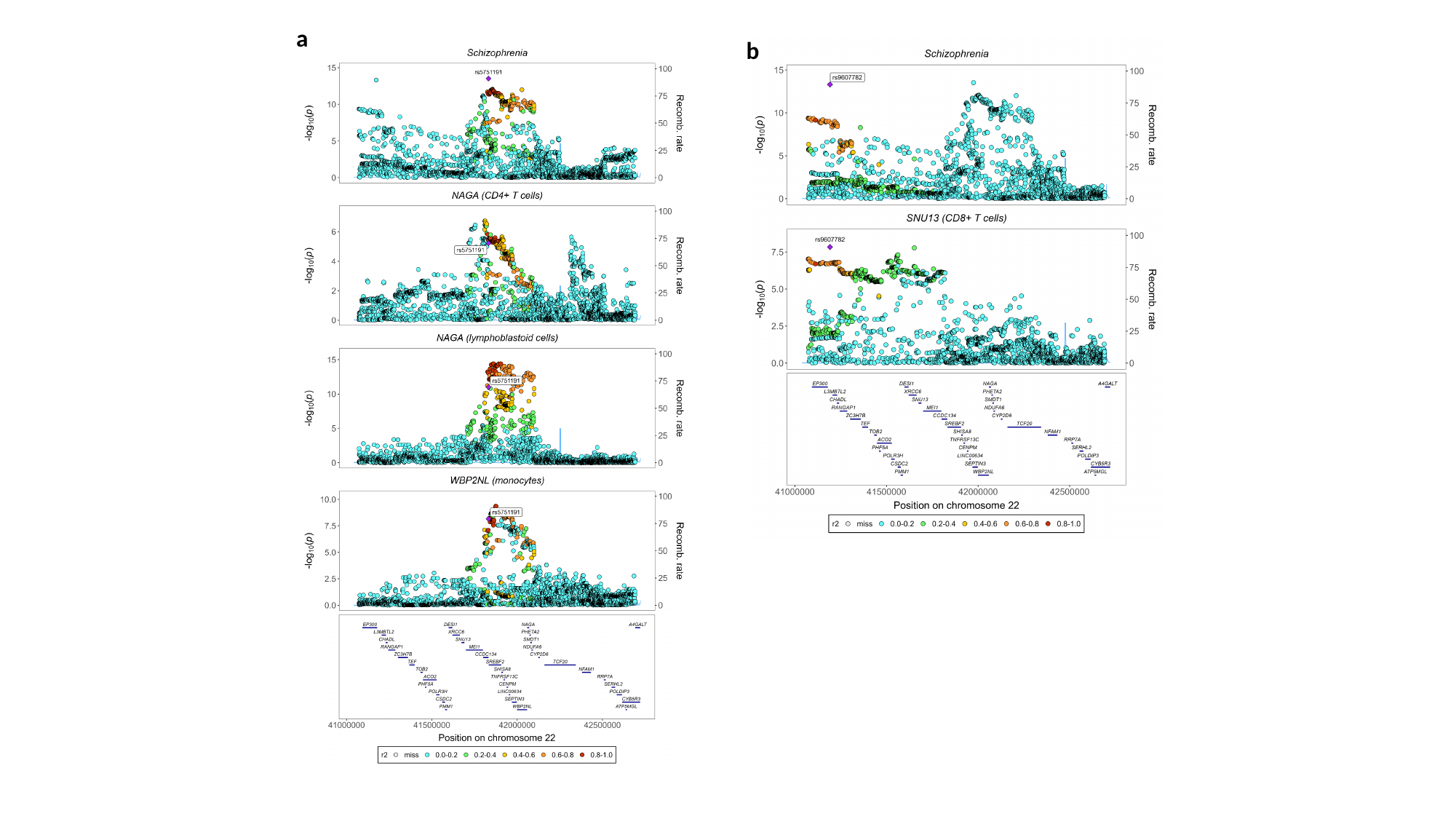

a
b

#### Slide 3
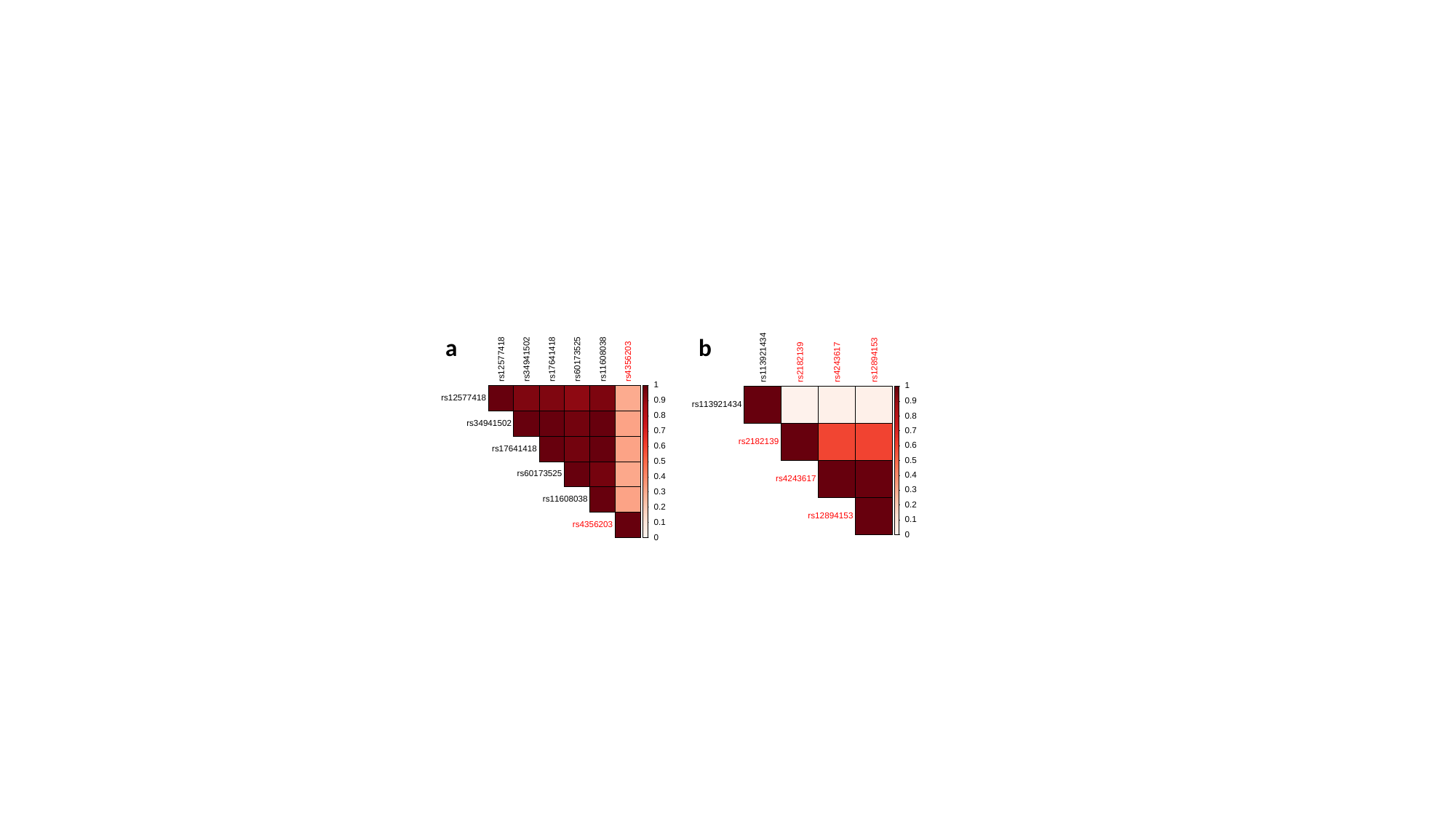

a
b

#### Slide 4
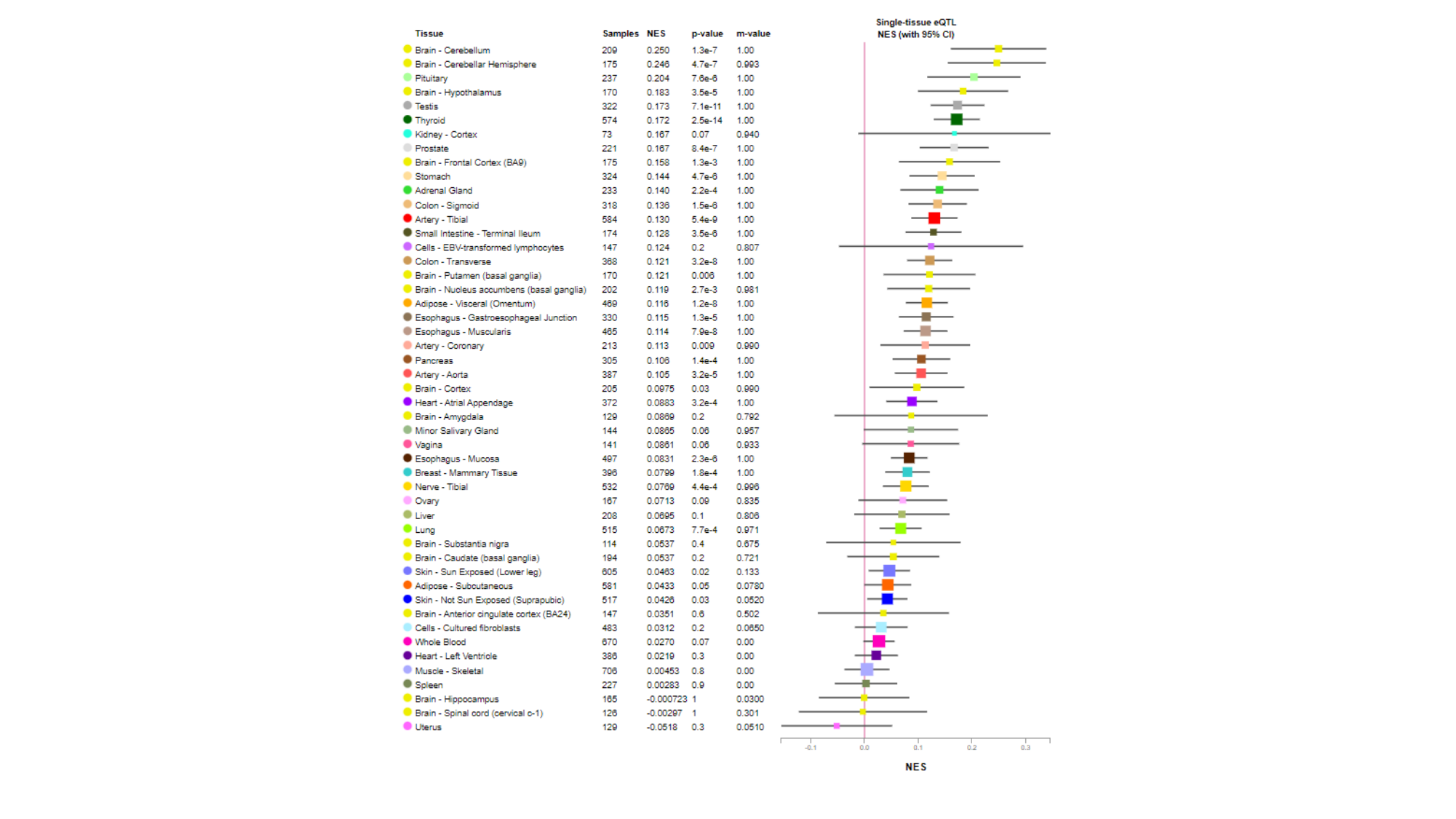
