## Supplementary Information for "A transcriptome-wide Mendelian randomization study in isolated human immune cells highlights risk genes involved in viral infections and potential drug repurposing opportunities for schizophrenia"

Stacey et al.

**SUPPLEMENTARY METHODS**

***Summary datasets.*** For this study we utilized publicly available summary data from: (i) the largest and most recent GWAS of SCZ available from the psychiatric genetics consortium (PGC) (<https://pgc.unc.edu/for-researchers/download-results/>) in up to 130,644 Europeans, including 53,386 cases; (ii) *cis*-eQTL studies of isolated human immune cell types available through the eQTL catalogue(1, 2) (<https://www.ebi.ac.uk/eqtl/>) (**Table 1**), as well as the genotype tissue expression (GTEx) project(3) (<https://gtexportal.org/home/>) and the eQTLgen (phase I) consortium(4) (<https://www.eqtlgen.org/phase1.html>); (iii) GWASes of autoimmune thyroid disease(5) (European; up to 30,234 cases, 725,172 controls), cognitive performance(6) (European; up to 257,841 participants; <https://ftp.ebi.ac.uk/pub/databases/gwas/summary_statistics/GCST006001-GCST007000/GCST006572/>), diabetes or endocrine disease(7) (mixed, mostly European; up to 51,949 cases, 432,649 controls; <https://www.ebi.ac.uk/gwas/publications/33959723>), hypothyroidism or myxedema(7) (mixed, mostly European; up to 23,497 cases, 461,101 controls; <https://www.ebi.ac.uk/gwas/publications/33959723>), thyroid problem (not cancer)(7) (mixed, mostly European; up to 28,254 cases, 456,344 controls; <https://www.ebi.ac.uk/gwas/publications/33959723>), and vitiligo(8) (European; up to 4,680 cases, 39,586 controls; <https://www.ebi.ac.uk/gwas/publications/27723757>).

To select datasets from the eQTL catalogue, we first identified those derived from isolated human immune cell types. After running TWMR analyses, we then removed datasets flagged by the generalised summary-data-based Mendelian randomization (GSMR)(9) software tool as having too many variants with more than a 20% difference (--diff-freq 0.2) in allele frequency compared to the LD reference panel used. This filtering step protected against large ancestral differences between the exposure and outcome samples and left us with 29 *cis*-eQTL datasets generated by 15 studies from 11 unique immune cell types for reporting and subsequent analyses.

***Statistical colocalization.*** We performed statistical colocalization analyses using two software tools: (i) the Coloc R package (v5.1.0.1)(10); and (ii) a C++ implementation of pair-wise conditional and colocalization (PWCoCo) analysis(11). In Coloc, we utilized the coloc.abf() function with priors and parameters set to default. The Coloc method has been described in detail previously, but briefly, it is a suite of Bayesian tools that takes locus-specific summary data for a pair of traits as input. The coloc.abf function returns posterior probabilities (PPs) to estimate the likelihood that five distinct scenarios – or hypotheses – are true, assuming a single causal variant for each trait. These five hypotheses are: H_0_, there is no causal variant for either trait at the locus; H_1_/H_2_, there is a causal variant for the first/second trait only; H_3_, there are two distinct causal variants at the locus, one for each trait; H_4_, both traits share the same causal variant. PWCoCo integrates Coloc with GCTA-COJO to enable colocalization after conditioning on independent regional signals. In PWCoCo, we set the *p*-value threshold for identifying significant signals in eQTL data from the eQTL catalogue and the GTEx project to the estimated empirical *p*-value for the corresponding eGene and eQTL dataset, as described above (***Transcriptome-wide Mendelian randomization***). Conversely, we set the *p*-value thresholds for SCZ and eQTLgen to *p*<5x10^-8^. For both Coloc and PWCoCo, we considered a H_4_ posterior probability (PP_H4_) >0.8 to be robust evidence of colocalization between two traits, and a PP_H4_>0.5 but ≤0.8 to be suggestive evidence though with potential for confounding by LD, as per Sun et al. (2018)(12). We generated regional association plots using the geni.plots R package (v0.1.0) using the INTERVAL reference genotype panel mentioned above(13).

***Enrichment analysis of regulatory data*.** To determine whether IRF3 ChIP-seq peaks were enriched at SCZ risk loci, we utilized the GWAS analysis of regulatory or functional information enrichment with LD correction (GARFIELD) v2 software tool(14), available at <https://www.ebi.ac.uk/birney-srv/GARFIELD/>. We downloaded all available human IRF3 ChIP-seq data from the encyclopedia of DNA elements (ENCODE) project, which originated from five distinct experiments: two using GM12878 cells (ENCSR000DZX, ENCSR408JQO) and one each using HeLa-S3 (ENCSR000EDF), HepG2 (ENCSR000EEJ), and SK-N-SH (ENCSR422ZAO) cells. To ensure we were inputting high quality and reproducible regulatory data, for each experiment we downloaded .bed files containing only the irreproducible discovery rate (IDR) thresholded peaks. We excluded the data for HepG2 cells due to an insufficient number of peaks relative to the other four datasets. We utilized the SCZ GWAS summary data from Trubestkoy et al. (2022)(15) and tested for enrichment using two different *p*-value thresholds to define SCZ risk loci: one set at genome-wide significance (*p*<5x10^-8^) and the other using a less stringent threshold of *p*<1x10^-5^. We used in-house R scripts (available on request) to format the IRF3 ChIP-seq peak and SCZ GWAS summary data ready for input to GARFIELD. We considered an annotation to be significantly enriched at SCZ risk loci by applying an adjusted *p*-value threshold (*p_adj_*<0.012) corrected for the number of effective IRF3 ChIP-seq annotations using the garfield-Meff-Padj.R script provided as part of the GARFIELD software. We extracted all variants overlapping an enriched IRF3 ChIP-seq annotations using the ‘garfield_extract_variants_overlapping_enriched_annotations.sh’ bash script. Finally, we generated a network graph depicting the overlapping loci and variants using Cytoscape v3.10.1.

**SUPPLEMENTARY RESULTS**

**Most of the validated schizophrenia-associated genes replicated in the whole blood *cis*-eQTL data from eQTLgen.**

In addition to validating our TWMR findings using statistical colocalization, we sought to replicate them using the whole blood *cis*-eQTL data from the eQTLgen consortium. We selected whole blood as a proxy tissue because most of the immune cells investigated in our discovery analyses should be represented within blood samples. We restricted replication analyses to the set of validated SCZ-associated genes. Overall, 51 out of 61 genes were present in the eQTLgen data and had at least one suitable instrument (see **Methods**), of which 41 replicated with *p_adj_*< 0.05/61 genes (**Fig. S1**, **Table S6**). Furthermore, of the 41 genes that replicated, 38 had directionally consistent estimates across both the immune cell and whole blood *cis*-eQTL datasets. These findings support the broad replicability of the validated SCZ-associated gene set.

**The schizophrenia-associated gene set contains mutationally constrained genes and genes associated with rare neurodevelopmental, neurological, and immunological disorders.**

Previous studies have shown that effector genes for neuropsychiatric disorders, including SCZ, tend to be: (i) mutationally constrained, (ii) causal genes for Mendelian or oligogenic forms of disease with neurodevelopmental or neurological symptoms, and (iii) preferentially expressed in the brain. We therefore annotated the validated SCZ gene set with these features. Overall, 12 of the 61 genes were mutationally constrained, as evidenced by extreme probabilities of loss of function intolerance (pLi>0.9) according to the gnomAD database (**Fig. 3**, **Table S7**). Furthermore, 15 of the 61 genes were annotated as being associated with at least one rare Mendelian or oligogenic form of disease according to either the OMIM or Orphanet database, of which 11 were associated with neurodevelopmental or neurological features (**Table S7**). Conversely, only four genes from the validated SCZ-associated gene set were annotated with brain-enriched expression (**Table** S7, see **Methods**).

Additionally, according to the OMIM and Orphanet databases, two of the genes from the validated gene set were associated with disorders involving immunological symptoms. *MAD1L1*, which encodes mitogen arrest deficient 1 like 1 protein, was associated with B cell lymphoma and mosaic variegated aneuploidy syndrome 7 with inflammation and tumour predisposition (**Table S7**), both of which are characterized by immune dysfunction. Furthermore, *IRF3*, which encodes interferon regulatory factor 3, was associated with herpes simplex virus-1 (HSV-1)-induced encephalitis (**Table S7**)(16, 17). HSV-1-induced encephalitis is characterized by acute inflammation in the brain and can present with psychotic symptoms(18, 19). Thus, not only does this indicate a clear link between *IRF3* and inflammation, but it also hints at a mechanism by which *IRF3* may contribute towards SCZ-related symptoms.

**Manual curation reveals three additional high confidence candidate effector genes unlikely to be confounded by horizontal pleiotropy.**

For the 11 bi- or multi-gene clumps, we sought to distinguish between genuine signals and those confounded by horizontal pleiotropy through manual curation. A clump on chromosome 1 contained two genes from the validated SCZ-associated gene set, one of which was the protein-coding gene *RERE* and the other was an overlapping non-coding antisense RNA (*RERE-AS1*) (**Fig. 3**), which, if biologically relevant, likely functions to regulate *RERE* expression. Similarly, another clump on chromosome 1 contained the protein coding gene *MAP7D1* and a long non-coding (lnc) RNA (ENSG00000285184) (**Fig. 3**). Since the biological support for the two non-protein coding genes was weaker relative to the protein-coding genes at both loci (i.e., poorer transcript support level, and lack of support across independent annotation databases; see **Methods**, **Table S8**), we prioritized the protein-coding genes *RERE* and *MAP7D1* as the most likely effector genes at these two clumps, respectively.

We also examined these 11 bi- or multi-gene clumps to assess whether any might contain distinct candidate effector genes for independent SCZ signals at the same locus. We therefore extracted those clumps containing at least two candidate effector genes that only colocalized with SCZ after conditional analyses (**Table S5**). This yielded two clumps (excluding the xHLA region) in totaL, both of which harboured two independent genome-wide significant (*p*<5x10^-8^) signals for SCZ(15). The first clump, located on chrosome 2, contained the candidate effector genes *FTCDNL1*, *TYW5*, and *MAIP1*, all of which colocalized with the same SCZ signal (indexed by rs1451488, not rs11680723) (**Table S5**). Conversely, the three candidate effector genes at the second clump, located on chromosome 22, colocalized with distinct SCZ signals, whereby *NAGA* and *WBP2NL* colocalized with a SCZ signal indexed by rs5751191 (**Fig. S3a**) while *SNU13* colocalized with the other nearby signal indexed by rs607782 (**Fig. S3b**) (**Table S5**). We therefore prioritized *SNU13* as an additional high confidence (i.e., no evidence of horizontal pleiotropy) candidate effector gene for SCZ.

***L3HYPDH*, but not *FOXN2* or *PIK3C2A*, is a candidate effector gene at a novel SCZ risk locus**

To determine whether any of the genetic variants used to instrument the three potentially novel candidate genes (*FOXN2*, *PIK3C2A*, *L3HYPDH*) in the present study had previously been associated with SCZ (*p*<5x10^-8^) in any other GWAS, we conducted single variant PheWASs using the Open Targets Genetics and the IEU OpenGWAS platforms (**Table S9**, **Table S10**). This search revealed that three correlated *FOXN2* instruments (rs72872744, rs79073127, rs79247094) had been highlighted in a previous SCZ GWAS by Bigdeli et al. (2021)(20), indicating that *FOXN2* did not reside at a novel SCZ risk locus.

To determine whether the two remaining genes, *PIK3C2A* and *L3HYPDH*, had previously been implicated in SCZ by any other nearby variants, we conducted gene-based look ups using the ‘locus-to-gene’ (L2G) pipeline from the Open Targets Genetics platform (see **Methods**). This search revealed that *PIK3C2A* was annotated by the L2G pipeline as a possible effector gene at a GWAS signal (indexed by rs4356203) comparing SCZ and bipolar disorder (BPD) cases vs. controls from Ruderfere et al. (2014)(21) (**Table S11**). We checked whether this variant was in LD with any of the five *PIK3C2A* instruments used in this study and found it to be in moderate LD with each of them (**Fig. S3a**). Thus, the *PIK3C2A* signals detected in our SCZ TWMR analyses was not independent of the previously observed SCZ/BPD GWAS signal.

We also found three previous SCZ-related signals(22-24) potentially attributable to *L3HYPDH* according to the L2G pipeline (**Table S11**), though the LD between these signals and the *L3HYPDH* instrument from this study (rs113921434) was negligible (**Fig. S3b**). Moreover, while the ‘variant-to-gene’ (V2G) pipeline from Open Targets assigned the *L3HYPDH* gene with high confidence to the *L3HYPDH* instrument used in this study, other nearby genes (e.g., *RTN1*, *CCDC175*) were favoured by the V2G pipeline as the most likely effectors at the previous SCZ-related GWAS signals at the locus (**Table S12**). Taken together, these findings indicate that the *L3HYPDH* signal detected in our SCZ TWMR analyses was independent of the previously observed SCZ-related GWAS signals, suggesting that *L3HYPDH* is a novel candidate effector gene.
